# Reference intervals of amino acid for newborns in North Asia: Diagnosing Inborn Errors of Metabolism Using Liquid Chromatography Tandem Mass Spectrometry

**DOI:** 10.1101/2021.04.13.21255444

**Authors:** T.A. Bairova, A.V. Belskikh, A.V. Atalyan, O.V. Bugun, N.V. Nemchinova, L.V. Rychkova

**Affiliations:** Laboratory of Personalized Medicine, Scientific Centre for Family Health and Human Reproduction Problems, Irkutsk, Russia; Functional Group of Information Systems and Biostatistics, Scientific Centre for Family Health and Human Reproduction Problems, Irkutsk, Russia; Scientific Centre for Family Health and Human Reproduction Problems, Irkutsk, Russia

**Keywords:** Inborn errors of metabolism, newborn screening, tandem mass spectrometry

## Abstract

**Aim:** To establish reference intervals (RI) for blood amino acids (AA) in healthy newborns of North Asia measured by liquid chromatography tandem mass spectrometry (LC-MS/MS) and evaluate their differences from respective reference values for newborns from other populations.

**Objectives:** A cross-sectional study of 381 healthy newborns was conducted. De-identified dried blood spots annotated by age, birth-weight, and sex were obtained from 381 healthy newborns aged 0–7 days. Data was collected from April to May of 2020.

**Methods:** Dried blood spots collected from filtered paper were used to analyze and measure of 13 derivatized amino acids using LC-MS/MS method. Nonparametric statistical approaches were used to generate 2.5th–97.5th percentile distributions for newborns in North Asia in accordance with CLSI EP28-A3c.

**Results:** Reference intervals (RI) for phenylalanine, tyrosine, citrulline, alanine, ornithine, proline in North Asian newborns differ slightly from those of newborns in other countries around the world. This allows for the use of universal RI in the diagnosis of congenital metabolic disorders of the indicated amino acids (AA) in different populations around the world. The RI for branched-chain essential AA (valine, leucine, and isoleucine are metabolic criteria for maple syrup disease) are higher in North Asian infants as compared to infants in other populations. In addition, the RI for arginine, aspartic acid, and glutamic acid in North Asian newborns are higher than in newborns in other countries around the world. In our study, the RI for methionine in newborns were lower than in many countries worldwide [McHugh D, 2011]. For optimal clinical practice, RI for certain AA in newborns (valine, leucine, isoleucine, methionine, arginine, aspartic acid, glutamic acid) should be determined for specific populations-further augmenting the diagnosis of inborn errors of metabolism.

## INTRODUCTION

Inborn errors of metabolism (IEM) are ‘rare genetic disorders which invariably lead to meta-bolic defects accompanied by an accumulation of toxic intermediate substances. The absence of specific clinical manifestation(s) determines the complexity of diagnostics. IEM as a disease can begin at any age, but often manifests right after birth. The consequences are dangerous and without appropriate therapy and nutritional limitations is oftentimes fatal.

Currently, more than 2000 cases of IEM have been identified and described. [1] Factors that influence the prevalence of the disorder(s) vary greatly amongst countries and social groups: 1:667 in Saudi Arabia [2], 1:784 in UK [3], 1:1200 in Canada [4], from 1:4100 [5] to 1:2900 [6] in Germany, from 1:1178 to 1:8304 [7] in China, 1:1944 in Egypt [8], 1:2396 in Portugal [9], 1:2800 in South Korea [10], 1:2916 in Malaysia [11], 1:3165 in Singapore [12], 1:5882 for Taiwan [13], 1:9000 in Japan [14]. Key factors include variations in analytical techniques [15], total number of IEMs analyzed, age differences at onset of disorder manifestation, and ethnicity and race of studied population(s) [16, 17]. Reference AA concentration ranges for 0-5 days of life were established in the R4S project, and included 45 countries [18]. This trial included 47 US states, Australia, several countries of the European Union and Africa. Japan, South Korea, Malaysia, and Taiwan represented the Asian territories involved in this project. There were no representative regions from Northern Asia. The possibility of using reference ranges, established for Europe, in the evaluation of AA levels in Northern Asia is not clear – herein lies the rationale for conducting the current study.

The aim of this study was to establish reference intervals for AA levels by LC-MS/MS, which have not been conducted in Northern Asia. Our secondary objective was to compare Reference Intervals (RI) with other parts of the world.

RI of 13 amino acids in newborns from North Asia were determined. The RI of the essential AA (valine, leucine, isoleucine, methionine, arginine) and nonessential AA (aspartic acid, glutamic acid) in newborns from North Asia differed from those in many nations around the world [McHugh D, 2011].

## 2. MATERIALS AND METHODS

### 2.1. Study design

Cross-sectional descriptive study.

### 2.2. Study population

400 dried de-identified blood spot (DBS) samples were collected in the regional perinatal center. Data for 19 samples were excluded due to extended collection date i.e., collection at 7 or further days of life, unusual bodyweight or unexpected concentrations. A possible reason for unexpected AA concentration levels could be explained by contamination or IEM. 381 DBS samples from healthy children were included in the analysis. Boys 203(53.28 %), girls–178 (46.72 %).

In accordance with the World Health Organization’s (WHO) criteria, «newborns» are categorically defined from the moment of birth to the 28th day of life [19]. The period from birth to the 7th day is considered «Early Neonatal». Newborn screening was conducted during this period for the purposes of early IEM detection.

Inclusion criteria:

1. Healthy male and female newborns;
2. Gestational age, 37-42 weeks;
3. Birth weight 2500 – 4000 g.;
4. APGAR score greater than 7 at 10;
5. Age 4-6 days.
6. Breastfeeding to the moment of sample collection.

Exclusion criteria:

1. Gestation less than 36 weeks;
2. Inborn errors of metabolism;
3. Positive family history with metabolic disorders;
4. Lethargy;
5. Irritation;
6. Poor feeding;
7. Tachypnea;
8. Persistent vomiting;
9. Abnormal movement;
10. Spasms;
11. Malformations;
12. Diagnosed disorders of the liver, kidneys, heart, skeletal muscles;
13. Prescribed drug therapy;
14. Mother’s diabetes mellitus disease.

The average age of infants that participated in our study was median (Me) (25th; 75th percentiles) 3 (3; 4) days. Mean birth body mass was M(25th;75th percentiles) 3408 (3095;3700)g, for boys M(25th;75th percentiles)3480 (3150;3730) g, for girls-3310 (3037,5;3637,5) g.

The study protocol was reviewed and approved by the local Ethic Committee of Scientific Centre for Family Health and Human Reproduction Problems (number 4.2; date of registration 12th of May 2020). All procedures were conducted in accordance with the Declaration of Helsinki. Prior to the collection of blood specimens, written consent was obtained from the parents of the newborns. Each newborn was assigned a unique identification (ID) number. Documented data collected from the mother and infants’ medical records included date of birth, sex, weight, height/length, and mother’s health.

### 2.3. Description of the study area

All mothers lived in the Irkutsk Region during pregnancy. The Irkutsk Region, located in the Asian part of Russia is 774 846 km^2^ (4,52 % of Russia). The territory’s continental climate type is characterized as arctic and/or sub-arctic (Fig. 1). Summer is cool. Winter is cold (down to -40 C). Average temperatures are below -10°C throughout the year.

**Fig 1.**
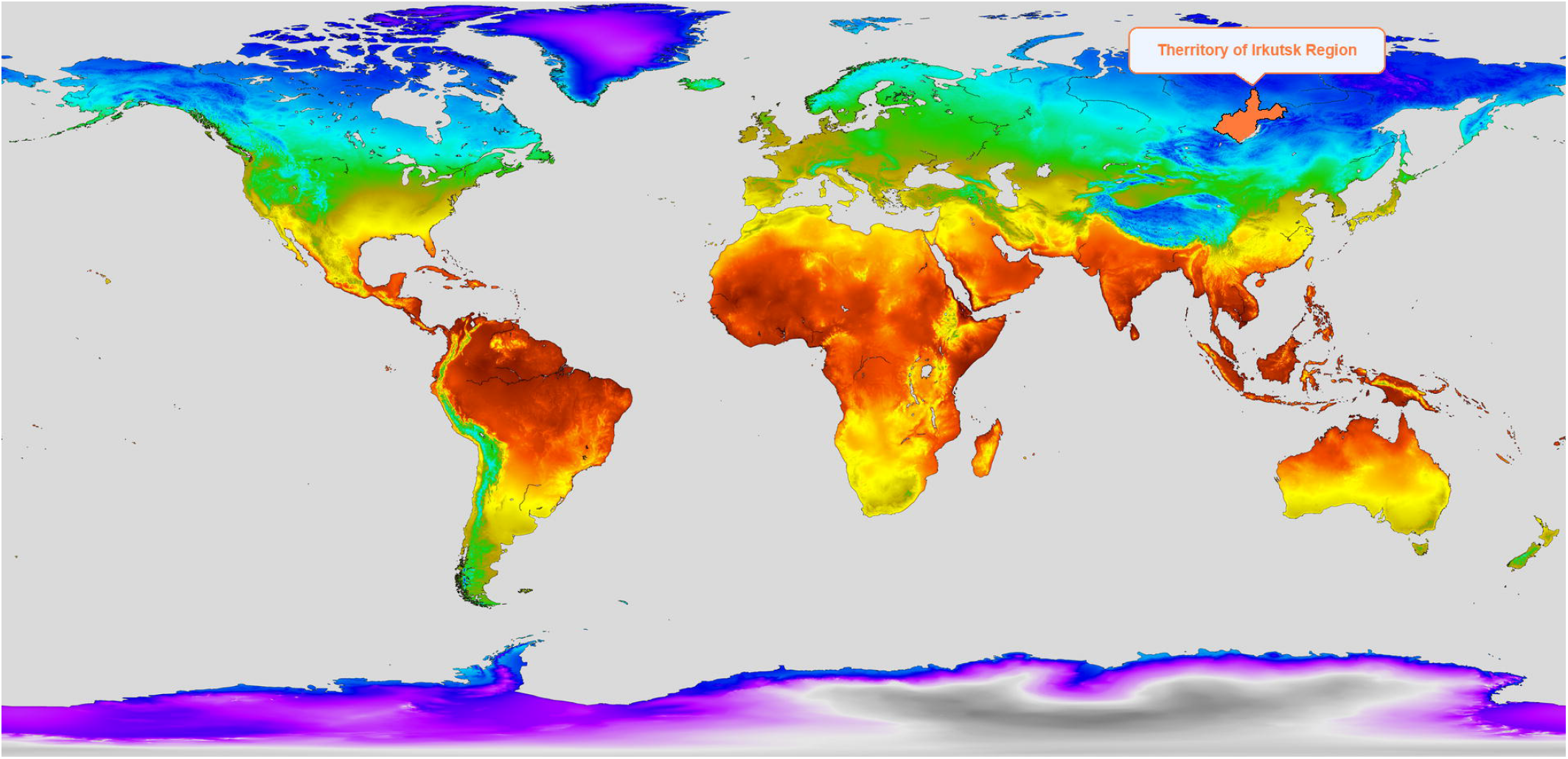
World map indicating the region in which the research was performed.

2. 39 million people live in the Irkutsk Region (actual for the 2020 year), with more than 30, 000 births registered annually. The population is multinational: Russians – 88%, Ukrainians – 1.2%, Tatars – 0.94%. Indigenous population consist of Buryats – 3.2%, evenks-0.05%, Yakuts – 0.04% and Tofalars – 0.03% (2010).

### 2.4. Sample Collection

Samples were collected, April-May of 2020. Blood samples were taken on the 4th day after birth in the regional perinatal center of Irkutsk Region, in accordance with the local newborn screening program. Blood specimens were collected by heel-stick and spotted on filter paper cards (Whatman 905, USA) a total of 5 spots, approximately 40 mkl of blood each. Filter paper cards were dried (DBSs) for 4 hours at room temperature on a horizontal, non-humid and nonabsorbent surface and then stored frozen at -20° C with a drying agent for 1-3 months.

### 2.5. Analysis of Amino Acids

Concentrations of 13 AA were evaluated: acid aspartic (Asp), acid glutamic (Glu), alanine (Ala), arginine (Arg), citrulline (Cyt), glycine (Gly), leucine (Leu) / isoleucine (Ile), metionine (Met), ornithine (Orn), phenylalanine (Phe), proline (Pro), tyrosine (Tyr), valine (Val). AA spectrum defined by reagent kit for AA level measurement “Amino Acid and Acylcarnitines from dried blood” (Chromsystems, Munich/Germany).

4 mm dried blood spot disks were punched out of the filter card and placed on to a 96-well microtiter plate. 200 mkl of the reconstituted internal standard was added. The plate was evaporated at 60°C and 600 rpm until dry. 60 mkl of derivatization reagent (butan-1-ol, n-butyl acetate, and hydrogen chloride) was added, the microtiter plate was sealed with a protective sheet, incubated for 15 min at 60°C and 600 rpm, and evaporated at 60°C and 600 rpm until dry. 100 mkl reconstitution buffer was added and agitated for 1 min at 600 rpm. 10 mkl was used for analysis with an optimized tandem mass spectrometer HPLC-MS/MS method. The analysis was performed with Shimadzu LCMS-8060 triple quadrupole mass spectrometer with Shimadzu LC-30AD HPLC system (Shimadzu, Kyoto, Japan). Optimization of the method was performed in accordance with the certified manufacturer’s specifications [20]. Optimal operating conditions included a 1.7-minute run time and mobile phase gradient with a flow rate of 20–200 mkl/min

### 2.6. Statistical Analysis

Statistical analysis was performed using STATISTICA (data analysis software system), version 10 (StatSoft, Inc.), and R – a free software environment for statistical computing and graphics (https://www.r-project.org/).

AA values, and characteristics of the newborns (gender, age, and weight) were described using descriptive statistics. Results were reported as median (Me) and 25, 75 percentiles for quantitative variables and percentages for the categorical variables. The nonparametric Mann–Whitney U test was used to analyze for differences in skewed distributed continuous variables in two gender groups. Empirical distribution were tested by the Kolmogorov-Smirnov test. Two-tailed α<0.05 were considered significant.

Moreover, 1, 2.5, 5, 10, 25, 75, 90, 97.5, 99, 99.5, 99.9 percentiles and the 95% confidence intervals were calculated for 13 AA. Bootstrap resampling was used to estimate confidence intervals (R packages: boot (), boot. ci()).

Calculation of sampling sufficiency was not done in accordance with requirements from Clinical and Laboratory Standards Institute (CLSI CA28-3, 2018). Normally, for RI estimation no less than 120 individuals are required.

## 3. RESULTS AND DISCUSSION

In this study, we calculated blood levels for 13 AA from DBS of healthy boys and girls in the early neonatal period (from birth to the 7th day of life) within North Asia (Table 1).

**Table 1.** Comparison of the medians Me (25; 75 percentiles) of AA in the blood of newborn boys and girls

| № | AA | All (n=381) | Boys (n=203) | Gerls (n=178) | p |
| --- | --- | --- | --- | --- | --- |
|  |  | Essential AA |  |  |  |
| 1 | Leu/Ile | 151.70 (113.93; 239.46) | 157.92 (112.10;256.69) | 148.75 (117.36;235.99) | 0.905 |
| 2 | Val | 89.53 (66.84; 116.38) | 89.95 (69.03;118.94) | 89.06 (65.82;114.36) | 0.322 |
| 3 | Met | 11.89 (9.23; 16.66) | 11.73 (9.08;16.95) | 12.06 (9.57;16.20) | 0.805 |
| 4 | Phe | 43.65 (34.80; 54.29) | 44.22 (35.25;54.83) | 42.97 (34.02;53.00) | 0.314 |
| 5 | Arg | 15.71(12.06; 22.41) | 16.36 (11.79;23.35) | 15.45 (12.08;22.18) | 0.500 |
|  |  | No-essential AA |  |  |  |
| 6 | Ala | 223.79 (144.10; 333.69) | 215.58 (130.37;333.69) | 225.66 (162.62;330.77) | 0.301 |
| 7 | Asp | 185.88 (120.33; 395.48) | 184.35 (122.76;397.55) | 186.51 (114.66;393.78) | 0.574 |
| 8 | Glu | 409.23 (337.50; 483.73) | 408.18 (320.99;485.87) | 410.48 (345.39;479.54) | 0.703 |
| 9 | Tyr | 62.75 (47.02; 86.18) | 62.61(47.65;86.25) | 62.80 (45.72;86.18) | 0.728 |
| 10 | Cyt | 9.13 (6.73; 11.56) | 9.38 (6.73;12.03) | 9.03 (6.73;11.29) | 0.628 |
| 11 | Gly | 346.00 (224.90; 412.30) | 343.64 (232.02;412.29) | 347.74 (221.66;411.24) | 0.583 |
| 12 | Orn | 77.33(58.34; 104.86) | 77.33 (58.34;107.63) | 77.44 (57.38;103.33) | 0.976 |
| 13 | Pro | 168.38 (127.09; 210.84) | 165.13 (124.99;210.20) | 171.99 (128.33;211.61) | 0.566 |

Amino acids (AA) were divided into two groups: essential and non-essential. Essential AAcannot be synthesized by a human organism and therefore must be obtained from food sources. In our trial, essential AA included valine, leucine, isoleucine, methionine, and phenylalanine. For newborns, arginine was also considered an essential AA.

When comparing AA levels, no significant differences were found between boys and girls. Similar results, without significant differences between sexes for newborns, were shown by Dietzen, D. J. (2016) within the US population [21], Jing Liu (2016) in the Chinese population [22] and Jaraspong Uaariyapanichkul (2018) in the Thai language [23]. Gender influences AA levels in older children and adults as opposed to newborns [24, 25]. Men have higher concentrations of essential AAs as compared to women [26].

A key factor in making a sound clinical diagnosis involves qualified interpretation of laboratory data, most notably the use of reference values for levels of AAs.. A few studies have examined AA reference intervals in newborns; however, in our study some tests were excluded from consideration due to the use of parametric statistical analysis, since our data did not show a normal distribution of values. Differences in statistical analysis were difficult to compare neonatal RIs across studies. Some experts considered the 1st percentile as the bottom range [18, 27, 30]. The upper bounds represented values from the 90 to 99.9 percentiles. This is why we calculated different percentiles for each AA (Table 2).

**Table 2.** Amino acid percentiles in neonatal dried blood spots 381 newborns, analyzed by tandem mass spectrometry (MS/MS)

| AA marker | Percentiles (mmol/L) and 95% CI |  |  |  |  |  |  |  |  |  |  |
| --- | --- | --- | --- | --- | --- | --- | --- | --- | --- | --- | --- |
|  | 1 | 2,5 | 5 | 10 | 25 | 75 | 90 | 97,5 | 99 | 99,5 | 99,9 |
| Essential AA |  |  |  |  |  |  |  |  |  |  |  |
| Leu/Ile | 68.1<br>(64.8, 77.5) | 77.3<br>(69.2, 84.8) | 85.1<br>(79.34, 89.46) | 91.6<br>(89.3, 96.9) | 112.4<br>(108.1, 118.6) | 227.8<br>(210.0, 261.0) | 357.6<br>(330.7, 394.2) | 495.0<br>(444.6, 577.9) | 594.9<br>(502.5, 725.5) | 654.1<br>(544.0, 923.5) | 844.0<br>(601.0, 923.5) |
| Val | 31.9<br>(25.7, 40.0) | 40.6<br>(36.6, 42.7) | 43.2<br>(40.9, 45.3) | 50.1<br>(45.6, 53.66) | 66.9<br>(64.8, 71.3) | 116.3<br>(111.9, 123.8) | 183.4<br>(160.6, 202.8) | 257.0<br>(232.6, 305.9) | 322.4<br>(265.7, 409.4) | 365.9<br>(280.2, 536.1) | 488.9<br>(314.4, 536.1) |
| Met | 4.7<br>(3.4, 5.6) | 5.6<br>(4.9, 6.3) | 6.4<br>(5.7, 7.0) | 7.24<br>(6.9, 7.6) | 9.2<br>(8.9, 9.6) | 16.3<br>(15.4, 17.1) | 21.9<br>(20.1, 24.6) | 30.0<br>(26.3, 34.0) | 34.2<br>(30.2, 38.4) | 36.3<br>(31.4, 39.0) | 38.8<br>(34.7, 39.0) |
| Phe | 19.1<br>(18.7, 22.6) | 22.6<br>(19.5, 24.8) | 25.1<br>(22.9, 26.7) | 28.4<br>(26.1, 29.8) | 35.2<br>(33.4, 36.9) | 54.3<br>(51.6, 563) | 63.9<br>(61.3, 67.0) | 82.4<br>(72.4, 91.4) | 92.6<br>(83.5, 121.2) | 98.0<br>(84.6, 127.9) | 124.4<br>(92.5, 127.9) |
| Arg | 4.9<br>(4.1, 5.7) | 5.7<br>(5.1, 7.1) | 7.2<br>(6.0, 7.9) | 8.7<br>(8.0, 10.0) | 12.2<br>(11.0, 12.8) | 22.5<br>(21.6, 24.0) | 30.8<br>(27.4, 35.3) | 46.8<br>(41.8, 53.6) | 56.4<br>(48.5, 69.8) | 66.0<br>(53.3, 76.2) | 73.<br>(59.7, 76.2) |
| No-essential AA |  |  |  |  |  |  |  |  |  |  |  |
| Ala | 55.4<br>(44.3, 61.1) | 61.1<br>(58.4, 69.9) | 71.9<br>(63.0, 83.0) | 96.3<br>(76.7, 109.1) | 143.2<br>(130.3, 158.6) | 335.3<br>(310.9, 362.0) | 416.3<br>(403.4, 433.5) | 483.0<br>(455.3, 525.6) | 540.3<br>(485.8, 1094.8) | 647.5<br>(504.7, 1436.6) | 1309.8<br>(548.0, 1437.0) |
| Asp | 69.5<br>(65.1, 75.6) | 75.5<br>(70.3, 79.6) | 81.2<br>(76.3, 85.6) | 90.7<br>(85.2, 95.7) | 119.9<br>(110.6, 125.9) | 383.8<br>(336.9, 431.8) | 553.8<br>(512.1, 604.5) | 755.8<br>(703.3, 880.8) | 902.3<br>(772.8, 1012.8) | 1007.5<br>(824, 1182.0) | 1119.4<br>(918.0, 1182.1) |
| Glu | 177.9<br>(150.7, 211.4) | 211.5<br>(187.1, 230.7) | 245.2<br>(215.9, 259.9) | 278.6<br>(258.5, 292.2) | 337.6<br>(316.3, 346.2) | 485.6<br>(465.9, 505.9) | 627.2<br>(565.9, 699.6) | 900.0<br>(767.7, 1083.2) | 1122.7<br>(932.0, 1460.1) | 1249.13<br>(1009.1, 1549.2) | 1516.14<br>(1157.0, 1549.0) |
| Tyr | 27.4<br>(24.1, 31.3) | 31.4<br>(27.8, 33.7) | 34.5<br>(32.0, 36.5) | 38.7<br>(35.9, 40.4) | 47.6<br>(45.0, 49.3) | 87.0<br>(80.1, 93.1) | 120.3<br>(105.2, 134.5) | 176.3<br>(161.6, 192.4) | 196.7<br>(179.6, 329.7) | 244.2<br>(185.7, 348.7) | 338.8<br>(201.2, 348.7) |
| Cyt | 3.9<br>(3.5, 4.4) | 4.4<br>(4.0, 4.6) | 4.6<br>(4.4, 5.0) | 5.4<br>(5.0, 5.6) | 6.7<br>(6.3, 7.2) | 11.5<br>(11.0, 12.1) | 14.2<br>(13.4, 15.2) | 18.4<br>(16.9, 20.2) | 20.4<br>(18.5, 27.4) | 20.9<br>(19.2, 43.8) | 35.7<br>(20.3, 43.9) |
| Gly | 97.6<br>(90.3, 130.7) | 132.5<br>(99.5, 145.7) | 150.9<br>(136.8, 159.0) | 176.8<br>(157.7, 186.3) | 224.7<br>(212.8, 243.0) | 411.9<br>(399.5, 438.1) | 492.1<br>(479.4, 521.7) | 593.9<br>(562.2, 663.6) | 665.4<br>(599.2, 771.3) | 697.5<br>(635.2, 807.6) | 794.1<br>(665.5, 807.6) |
| Orn | 32.2<br>(28.9, 37.0) | 36.9<br>(32.5, 39.5) | 40.3<br>(37.3, 43.7) | 46.6<br>(43.3, 49.1) | 58.5<br>(55.8, 61.1) | 104.3<br>(96.6, 113.5) | 136.4<br>(130.4, 149.6) | 200.0<br>(173.3, 243.9) | 249.9<br>(201.8, 276.6) | 266.1<br>(210.9, 406.6) | 358.3<br>(252.4, 406.6) |
| Pro | 64.1<br>(51.3, 70.9) | 70.7<br>(65.3, 85.3) | 85.9<br>(73.6, 92.5) | 97.2<br>(91.4, 103.0) | 125.9<br>(116.5, 131.4) | 208.4<br>(200.4, 220.7) | 271.2<br>(248.0, 289.6) | 356.6<br>(333.9, 429.9) | 441.1<br>(361.0, 476.9) | 449.6<br>(378.9, 520.1) | 497.53<br>(433.8, 520.1) |

At the next stage, we compared the RI of AA in newborns from North Asia with similar indicators in newborns among other populations. All studies presented used DBS samples and HPLC-MS / MS. Percentiles of the reference values from other studies were compared with 95% confidence intervals of the same percentiles calculated in our study sample of newborns. The decision on the comparability of the reference values was based on whether the value of the compared percentile was included in the calculated 95% confidence interval of our references (Table 3).

**Table 3.**
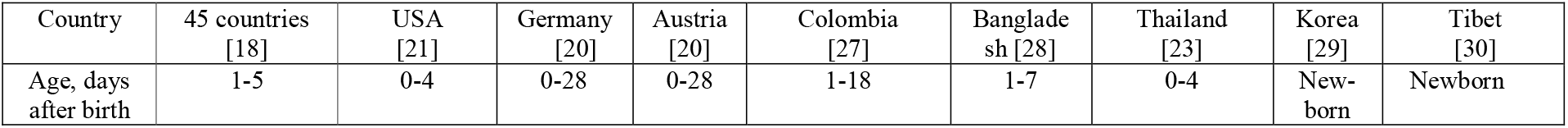
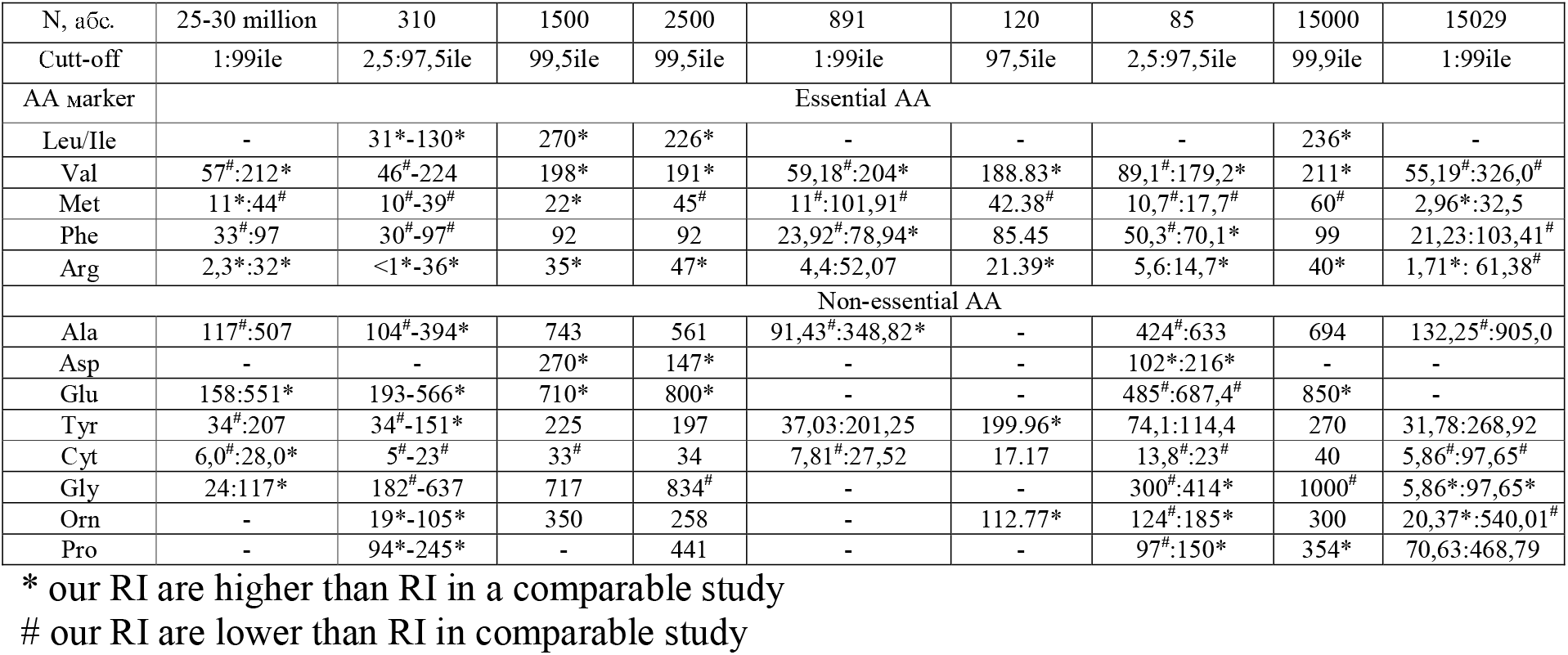
Comparison of the results of our studies of the AA content in the blood of newborns (0-7 days, N = 381) by tandem MS / MS mass spectrometry with the data of other researchers

Results from a comparative analysis of the AA reference values in newborns indicate the variability of the AA spectrum in newborns in different countries around the world. The RI analysis of essential AAs indicates that the RI of phenylalanine in newborns from North Asia is comparable with newborns from other countries. Despite the statistical significance of some of the differences, these differences are not large enough to indicate the use of different reference intervals for the diagnosis of phenylketonuria in clinical practice.

Differences in references for other essential amino acids were revealed. Our data indicate high concentrations of branched-chain AA (valine, leucine, and isoleucine) in newborns from North Asia as compared to other populations. An exception were the newborns from Tibet, whose concentrations of branched-chain AA were higher than those of the newborns in our study (Ch. Zhang, 2019) [30]. Disturbances in the metabolism of leucine, isoleucine and valine have been reported in maple syrup urine disease (MSUD). This is a rare (orphan) disease. The prevalence is 1: 110,000 newborns in Sweden [31] to 1: 1,148,000 in Korea [32]. MSUD diagnostics includes a quantitative assessment of the levels of leucine, isoleucine and valine in the blood with an assessment of the leucine / alanine ratio and genetic diagnostics with the detection of mutations in the branched-chain AA dehydrogenase genes: *BCKDHA, BCKDHB, DBT, DLD*. Moreover, the spectrum of mutations in these genes in Russian patients with MSUD differs from other populations [33].

The same pattern was found for arginine, aspartic acid, glutamic acid in which levels were higher than mentioned in other epidemiological trials [18, 20, 21, 23, 28, 29].

We found low levels of the essential amino acid methionine in North Asian babies as compared to babies in other populations around the world. It is well-known and widely accepted that essential AAs cannot be synthesized in humans, including newborns. Levels of AA are determined by placental circulation and breast milk [34]. In our trial, data specific to protein consumption thru breastfeeding was not collected. Yet, earlier trials show that high protein consumption can define AA metabolism for the first 4 months of life [35]. Respectively, AA blood levels in newborns directly correlate with AA metabolism in their respective mothers. As aforementioned, essential AA is not synthesized by the human body but rather sourced through food-intake; therefore, AA levels are influenced by nutritional patterns and thereby depend on cultural traditions, the availability and accessibility to food, presence of specific genes, (which assist in the metabolism of certain food products), and technological achievements.

North Asia is a territory of temperate to cold climate. Nutrition is a very important aspect in response to adaptation to the climate of certain geographical locations. Quantity and quality of meals should meet biological needs. Cold stress drives the formation of livestock economies, leading to high-caloric diets with a high content of animal-origin proteins and fats. Nutrition patterns define the metabolism of proteins and AA. Meat-eaters have 47% greater AA concentrations as compared with vegans (a diet that consist of little to no animal products and/or by-products); with significant differences in lysine, methionine, tryptophan, alanine glycine, and tyrosine levels [36, 37]. Furthermore, it has been demonstrated that diets high in carbohydrates did not lead to significant changes in AA levels; however, diets high in fat lead to increased levels of branched-chain AA: valine, leucine, and isoleucine [38]. Yet AA levels for North Asian newborns were higher than in other populations and races, excluding Tibet. J.A. Schmidt et al discovered another pattern during their clinical trial of 65,000 UK volunteers. They concluded that only a weak correlation existed between nutrition patterns and AA levels for meat-eaters, vegetarians, vegans, and fish-eaters i.e., pescatarians. However, repeated AA level evaluation shows differences in alanine and glycine levels - these levels were lower for meat-eaters as compared with other groups.

Analysis of the RI of non-essential AA (tyrosine, citrulline, alanine, ornithine, proline) shows that the RI in newborns from North Asia is comparable to newborns from other countries. Despite the statistical significance of some of the differences, these differences were not large enough to indicate the use of different RI for diagnosis within clinical practice. Essential AA level differences should be noted for Caucasian newborns from North Asia and newborns from Thai, due to higher levels in the Caucasian group. In 2006, Tan IK, et.al. noted such differences between Caucasian and Asian newborns [16]. At the same time, we found comparability of RI for essential AA for Caucasian newborns from North Asia and newborns from Tibet – there were no significant differences between groups in levels of valine, methionine, phenylalanine, and arginine.

## 4. CONCLUSION

Our work evaluated RI for 13 AA in newborns from North Asia, considering 2.5 and 97.5 percentiles as lower and upper limits of reference intervals. RI for AA are variable in different populations. The RI for phenylalanine, tyrosine, citrulline, alanine, ornithine, proline in North Asian newborns differ slightly from the RI for newborns in other countries around the world. This allows for the use of universal RI in the diagnosis of congenital metabolic disorders of the indicated AA in different populations around the world. The RI for branched-chain essential AA (valine, leucine, and isoleucine are metabolic criteria for maple syrup urine disease) are higher in North Asian infants than in infants in other populations around the world. In addition, the RI for arginine, aspartic acid, glutamic acid in North Asian newborns were higher than in newborns in other countries around the world. The RI for methionine in newborns within our study were lower than in many countries around the world. For optimal clinical practice, RI for certain AA in newborns (valine, leucine, isoleucine, methionine, arginine, aspartic acid, glutamic acid) should be determined for specific populations. This will help diagnose inborn errors of metabolism.

In an effort to increase the effectiveness of newborn screening for IEM expedient a unified approach for RI estimation, in accordance with «Defining, Establishing, and Verifying Reference Intervals in the Clinical Laboratory, Approved Guideline – Third Edition» (2010), where RI are considered at intervals between 2.5th and 97.5th percentiles.

A differential approach for AA RI is necessary. It allows for the optimization of analysis in IEM diagnosis, formation of a solution attraction algorithm, and harmonizes approaches to the formulation of regional IEM lists for each world population, with specific consideration given to race and ethnic differences.

## Supporting information

RAW data

## Data Availability

Authors declare that all data mentioned in current paper are fully available without restriction.

## Acknowledgement

The authors gratefully acknowledge the staff of the Regional Perinatal Center (head MD, Ph.D. NV. Protopopova). Department of Pathology for help in collecting material.

